# Increased plasma heparanase activity in COVID-19 patients

**DOI:** 10.1101/2020.06.12.20129304

**Authors:** Baranca Buijsers, Cansu Yanginlar, Aline de Nooijer, Inge Grondman, Marissa L. Maciej-Hulme, Inge Jonkman, Nico A.F. Janssen, Nils Rother, Mark de Graaf, Peter Pickkers, Matthijs Kox, Leo A.B. Joosten, Tom Nijenhuis, Mihai G. Netea, Luuk Hilbrands, Frank L. van de Veerdonk, Raphaël Duivenvoorden, Quirijn de Mast, Johan van der Vlag

**Author notes:** Corresponding author: Prof. Dr. Johan van der Vlag, Department of Nephrology (480), Radboud Institute for Molecular Life Sciences, Radboud university medical center, Geert Grooteplein 10, 6525 GA Nijmegen, The Netherlands. co-first (Baranca Buijsers, Cansu Yanginlar). co-second (Aline de Nooijer, Inge Grondman).

## Abstract

Reports suggest a role of endothelial dysfunction and loss of endothelial barrier function in COVID-19. It is well established that the endothelial glycocalyx-degrading enzyme heparanase contributes to vascular leakage and inflammation. Low molecular weight heparins (LMWH) serve as an inhibitor of heparanase. We hypothesize that heparanase contributes to the pathogenesis of COVID-19, and that heparanase may be inhibited by LMWH. Heparanase activity and heparan sulfate levels were measured in plasma of healthy controls (n=10) and COVID-19 patients (n=48).

Plasma heparanase activity and heparan sulfate levels were significantly elevated in COVID-19 patients. Heparanase activity associated with disease severity including the need for intensive care and mechanical ventilation, lactate dehydrogenase levels and creatinine levels. Use of prophylactic LMWH in non-ICU patients was associated with a reduced heparanase activity. Since there is no other clinically applied heparanase inhibitor currently available, therapeutic treatment of COVID-19 patients with low molecular weight heparins should be explored.

## Introduction

The coronavirus disease-2019 (COVID-19) pandemic is caused by the severe acute respiratory syndrome coronavirus 2 (SARS-CoV-2).^1^ Severe COVID-19 usually manifests as pneumonitis or acute respiratory distress syndrome (ARDS).^2,3^ Studies showed that upon hospital admission 59% of COVID-19 patients had proteinuria^4^, and 22% of the non-ventilated patients and 90% of the ventilated patients developed acute kidney injury (AKI).^5,6^ Endothelial barrier function is crucial in the regulation of fluid and protein extravasation, particularly in the lungs^7,8^ and in the kidneys.^9,10^ An important role for endothelial cell dysfunction in the pathogenesis of the complications of COVID-19 has been proposed by several studies.^11,12^ As pulmonary edema occurs when fluid leaks into alveoli, dysfunction of the endothelium is likely to contribute to pulmonary edema in COVID-19. Furthermore, it has been well established that proteinuria occurs when the endothelial barrier function in the glomerulus is compromised.^9,10,13^

Endothelial cells are covered with a thick layer of negatively charged glycosaminoglycans (GAGs), termed the glycocalyx. Heparan sulfate (HS) is the predominant sulfated GAG in the glycocalyx. HS contributes to the endothelial charge-dependent barrier function due to its negative charge.^14^ Degradation of HS by heparanase (HPSE), the only known mammalian HS-degrading enzyme, disrupts the endothelial glycocalyx and subsequent loss of endothelial barrier function, as observed in ARDS and proteinuric kidney diseases.^7,15–17^ In addition to compromising barrier function, HPSE generates a pro-inflammatory glycocalyx that promotes the binding of chemokines, cytokines and leukocytes to the endothelial cell surface. Inhibition of HPSE and/or HPSE deficiency is beneficial in experimental lung and kidney diseases.^7,15–18^ Notably, heparins and low molecular weight heparin (LMWH) that have been suggested to be beneficial for COVID-19 patients^19^, are potent inhibitors of HPSE activity.^20,21^

Taken together, we hypothesize that increased HPSE activity is one of the driving forces in severe COVID-19 manifestation and that HPSE may be inhibited by the use of LMWH in COVID-19.

## Results

### Plasma HPSE activity is elevated in COVID-19 patients

Demographics and baseline characteristics of COVID-19 patients are listed in Table 1. Several (experimental) disease models have shown that increased HPSE activity can lead to endothelial barrier dysfunction, which may be involved in the development of ARDS and proteinuria/AKI.^7,22,23^ Measurement of plasma HPSE activity levels in COVID-19 patients and healthy controls revealed that HPSE activity was significantly elevated in COVID-19 patients compared to healthy controls (Figure 1A). In line with the increased HPSE activity, HS plasma levels were also significantly elevated in COVID-19 patients compared to healthy controls (Figure 1B). Overall, these results suggest that SARS-CoV-2 infection causes an increase in the activity of HPSE in plasma and an increase in circulating HS.

**Table 1.**
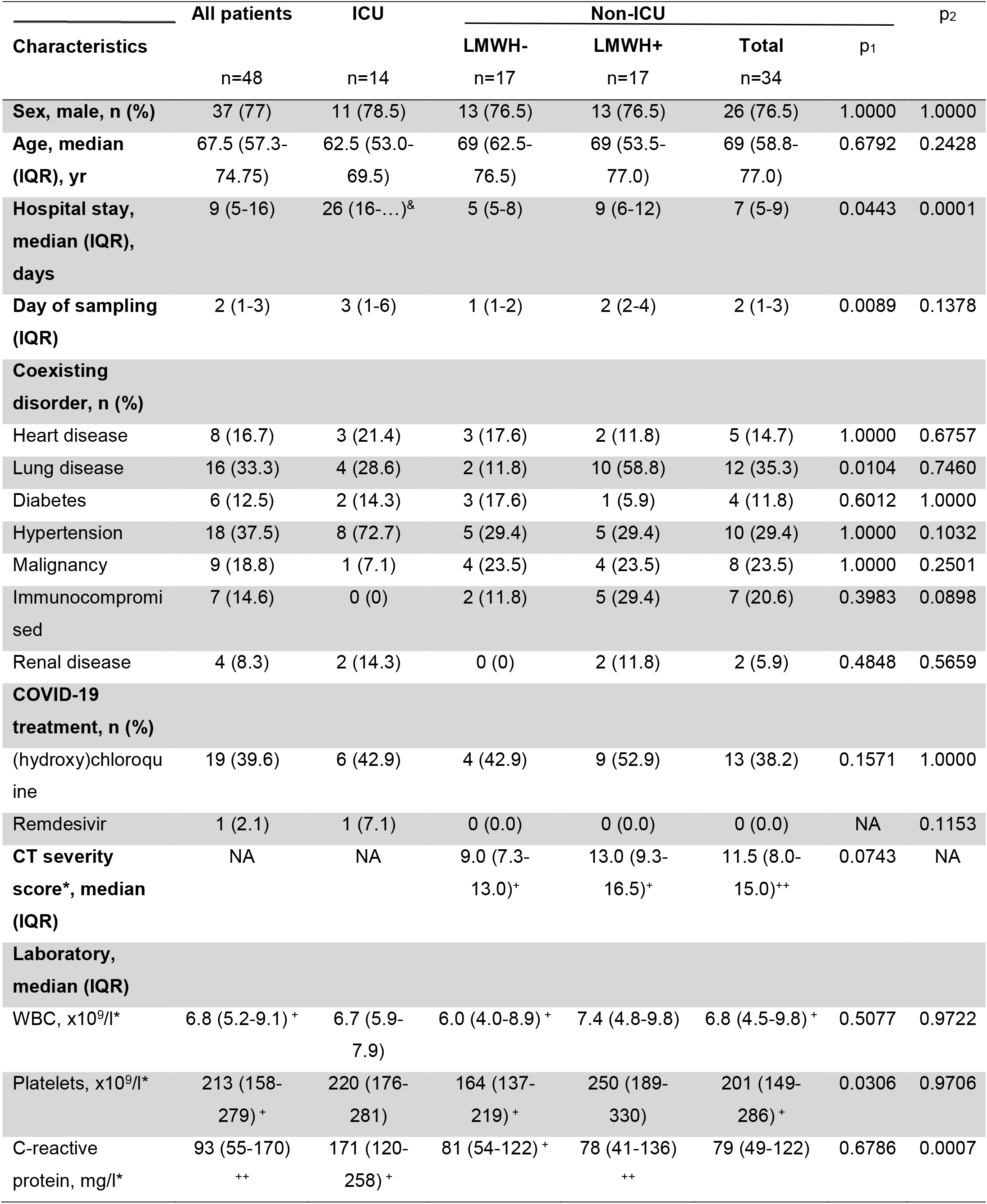
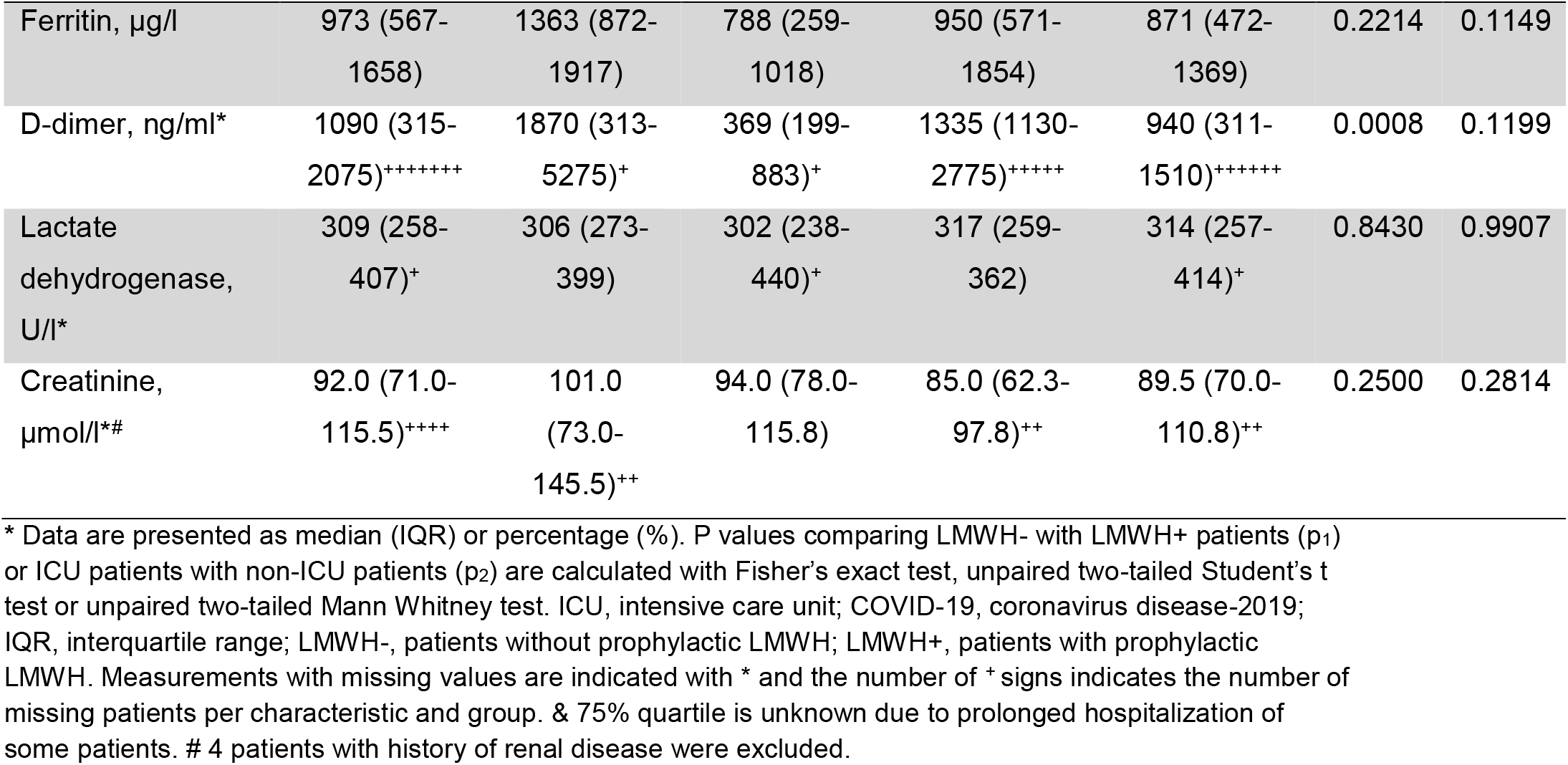
Demographics and baseline characteristic of COVID-19 patients*.

**Figure 1.**
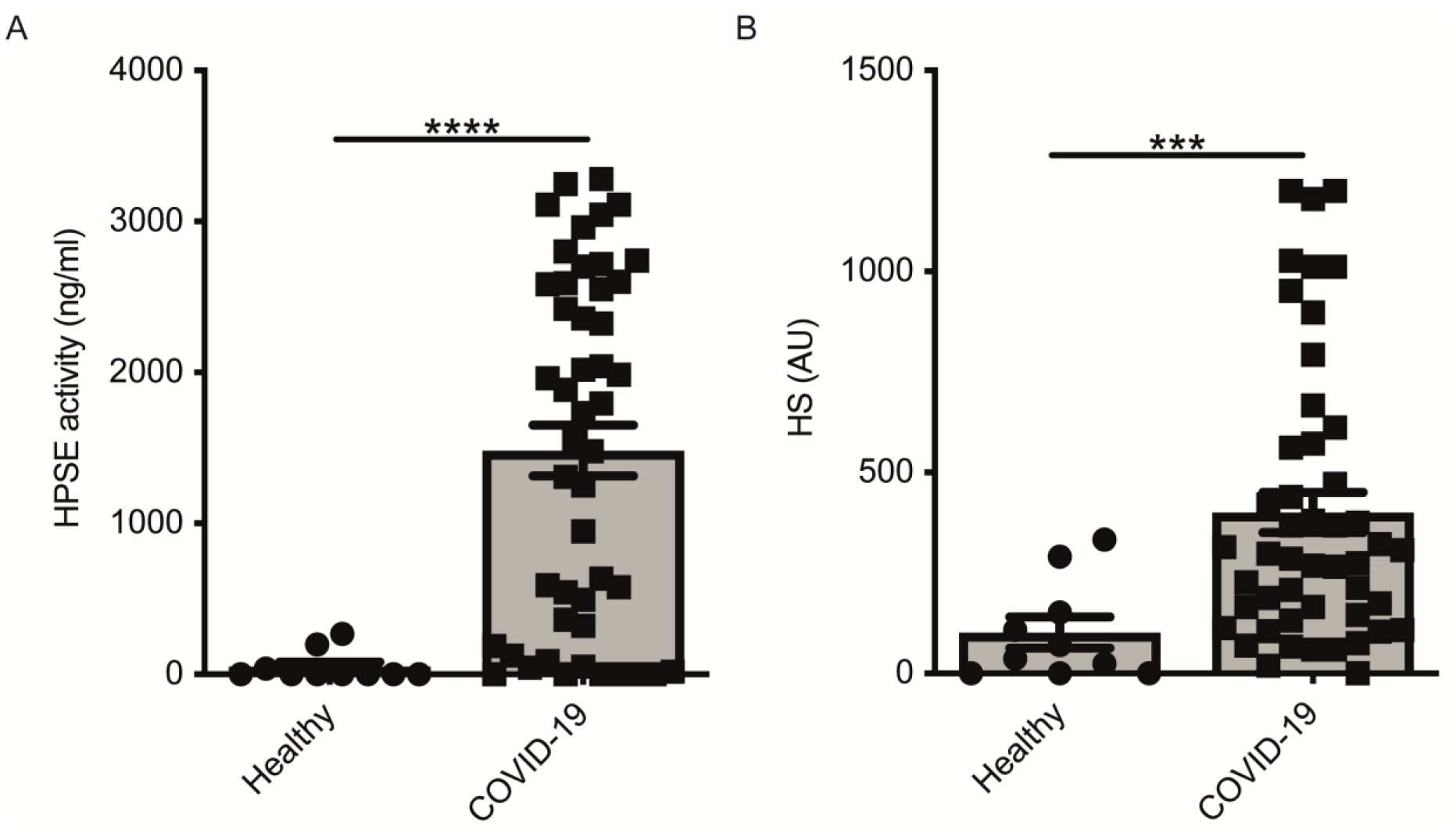
COVID-19 patients display increased HPSE activity and elevated levels of heparan sulfate in plasma. **A**. HPSE activity was increased in plasma of COVID-19 patients compared to healthy controls. HPSE activity was measured using an in-house developed ELISA with a specific anti-HS antibody. **B**. HS levels were increased in plasma of COVID-19 patients compared to healthy controls. HS levels were measured by an in-house developed competition ELISA using a specific anti-HS antibody. Data were presented as mean±SEM and tested for normal distribution with D’Agostino & Pearson omnibus normality test and statistical differences were calculated using Mann Whitney test (n=10 healthy; n=48 COVID-19, *** p<0.001, **** p <0.0001). HPSE, heparanase; HS, heparan sulfate; Healthy, healthy controls; COVID-19, coronavirus disease-19 patients; AU, arbitrary units.

### HPSE activity associates with COVID-19 disease severity

Next, we investigated whether HPSE activity levels were associated with COVID-19 disease severity. Plasma HPSE activity was significantly increased in both non-ICU and ICU patients compared to healthy controls, and HPSE levels in ICU patients were higher than in non-ICU patients (Figure 2A). Moreover, HS levels in plasma were also higher in both non-ICU and ICU patients compared to healthy controls (Figure 2B). Finally, plasma HPSE activity was significantly higher in patients in need of mechanical ventilation (Figure 2C), in patients with elevated LDH values (Figure 2D), and in patients with elevated serum creatinine values (Figure 2E). These findings reveal that patients with severe COVID-19 disease display higher plasma HPSE activity levels than patients with moderate COVID-19 disease.

**Figure 2.**
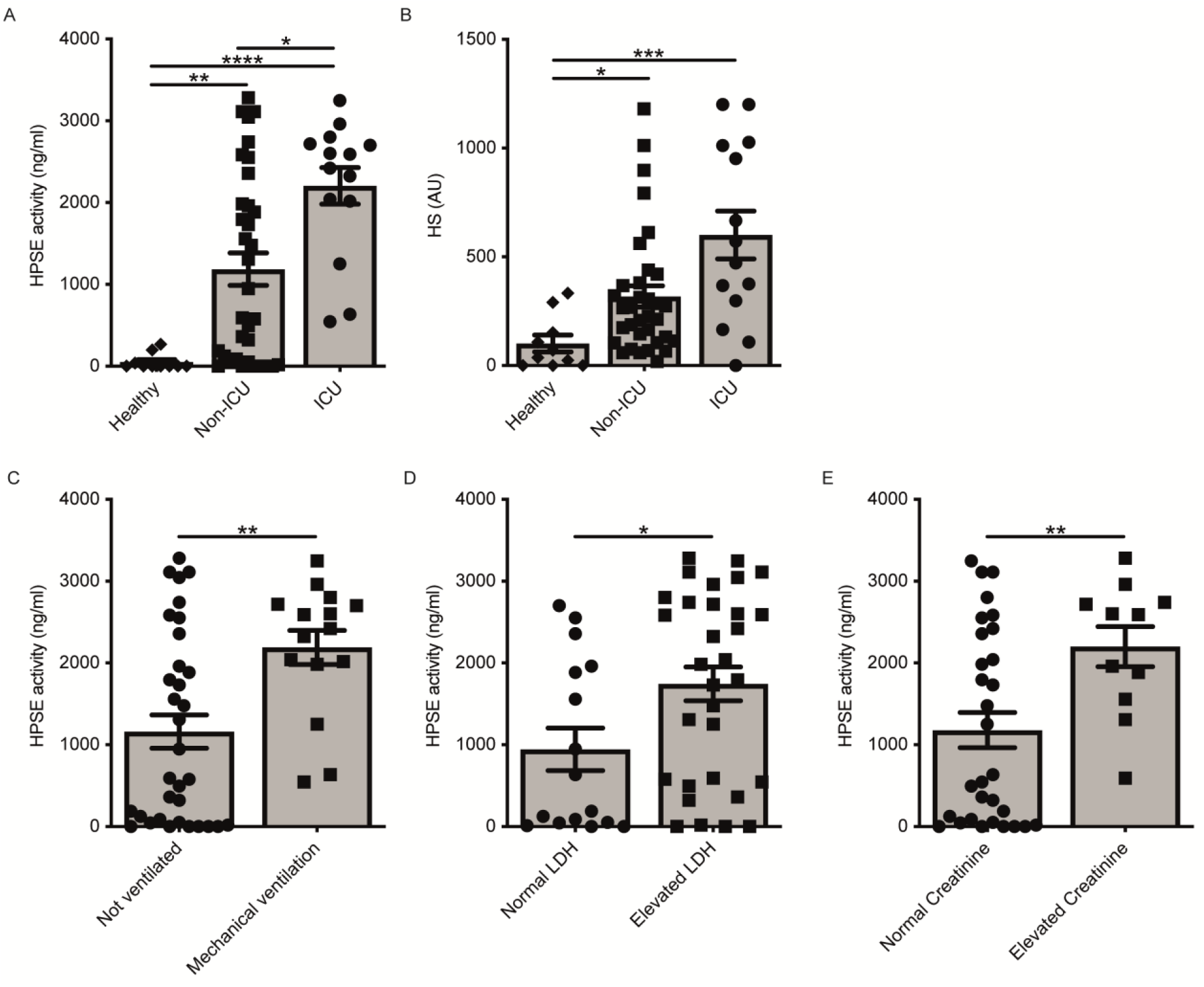
Increased plasma HPSE activity associates with COVID-19 disease severity. **A**. Plasma HPSE activity was significantly higher in ICU and non-ICU patients compared to healthy controls, and higher in ICU patients compared to non-ICU patients. (n=10 healthy; n=34 non-ICU; n=14 ICU) **B**. HS levels were significantly increased in plasma of ICU and non-ICU patients compared to healthy controls (n=10 healthy; n=34 non-ICU; n=14 ICU) **C**. HPSE activity was significantly higher in plasma of patients with mechanical ventilation compared to patients without mechanical ventilation. (n=33 not ventilated; n=15 mechanical ventilation) **D**. HPSE activity was significantly higher in plasma of patients with elevated LDH (>280 U/l) values compared to patients with normal LDH levels. (n=15 normal LDH; n=26 elevated LDH). **E**. HPSE activity was significantly higher in plasma of patients with elevated creatinine (>110 µmol/ for men and >90 µmol/ for women) values compared to patients with normal creatinine values. (n=30 normal creatinine; n=11 elevated creatinine; patients with history of renal disease were excluded from this analysis). HPSE activity was measured using an in-house developed ELISA with a specific anti-HS antibody. Data were presented as mean±SEM and tested for normal distribution with D’Agostino & Pearson omnibus normality test and statistical differences were calculated using Kruskal Wallis test followed by Dunn’s multiple comparison test, unpaired one-tailed Student’s t-test or unpaired one-tailed Mann Whitney test (* p<0.05, ** p<0.01, *** p<0.001, **** p<0.0001). HPSE, heparanase; HS, heparan sulfate; LDH, lactate dehydrogenase; Healthy, healthy controls; non-ICU: COVID-19 patients in normal hospital ward; ICU, COVID-19 patients in ICU; AU, arbitrary units.

### Use of LMWH is associated with lower HPSE activity in plasma of COVID-19 patients

Prophylactic treatment with LMWH is recommended for patients hospitalized with COVID-19^24^, whereas some experts recommend higher doses for critically ill patients. As LMWH inhibits HPSE activity, we analyzed the effect of prophylactic LMWH on HPSE activity in plasma of COVID-19 patients. Markedly, non-ICU patients who received LMWH displayed significantly lower plasma HPSE activity compared to non-ICU patients without LMWH prophylaxis (Figure 3A). According to literature, a single injection of 5000 units dalteparin would result in an estimated concentration of around 0.37 U/ml *in vivo*.^25^ We found a dose dependent inhibition of recombinant HPSE at concentrations between 0.0025 and 0.05 U/ml and full inhibition starting from 0.25 U/ml dalteparin *in vitro* (Figure 3B). These data suggest that the applied prophylactic LMWH dose is already effective in inhibition of HPSE activity within plasma of moderately diseased, but not severely ill, COVID-19 patients.

**Figure 3.**
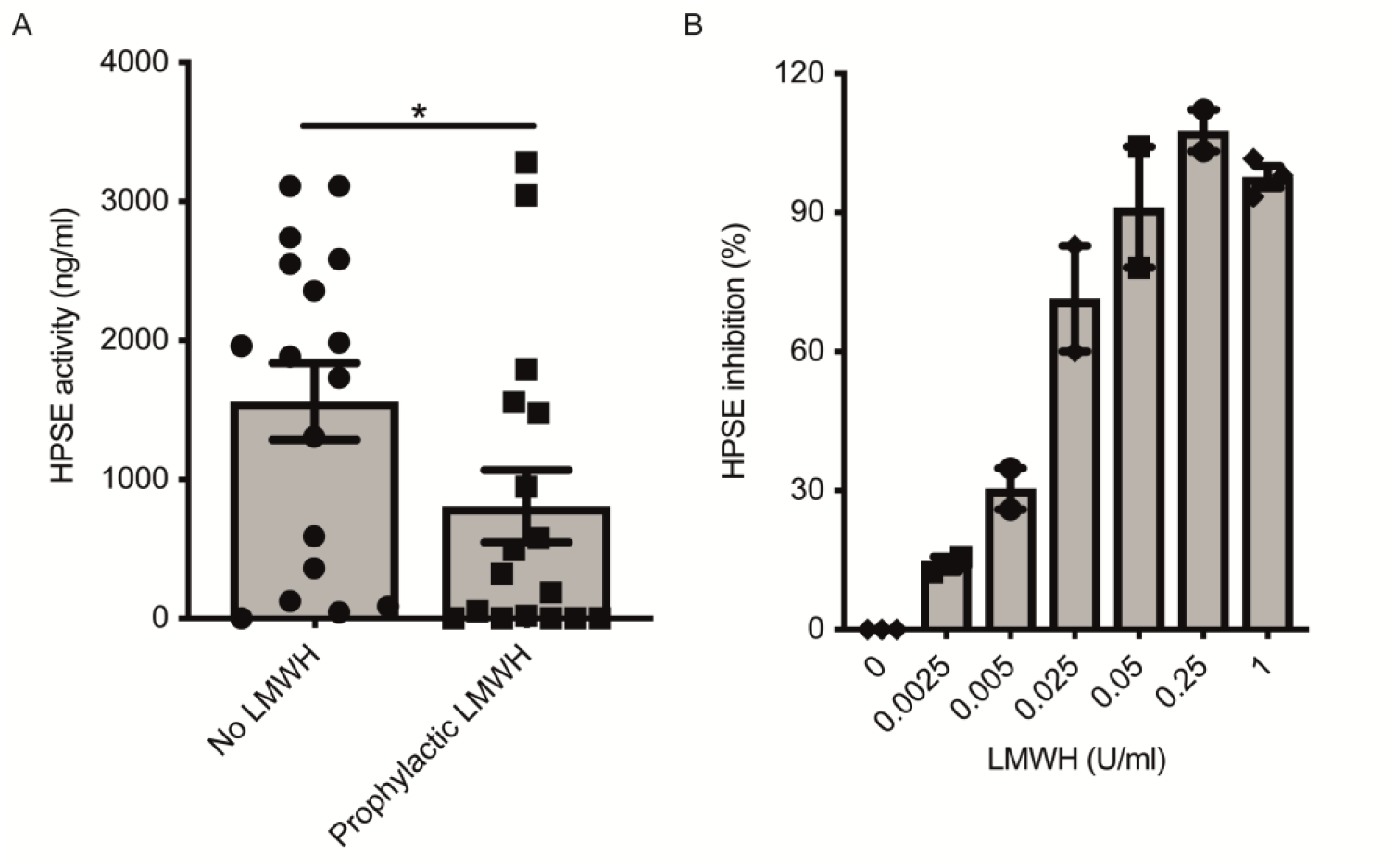
LMWH reduces plasma HPSE activity in moderately diseased COVID-19 patients. **A**. LMWH reduces HPSE activity in plasma of non-ICU patients with COVID-19, which was measured using in house developed HPSE activity assay. (n=17 for both groups, p<0.05). Data were presented as mean±SEM and tested for normal distribution with D’Agostino & Pearson omnibus normality test and statistical difference was calculated using unpaired one-tailed Mann Whitney test. **B**. LMWH inhibits recombinant HPSE activity *in vitro* in a dose dependent manner. HPSE activity was measured using an in-house developed ELISA with a specific anti-HS antibody. (n=3) HPSE, heparanase; LMWH, low molecular weight heparin.

## Discussion

COVID-19 appears to be a disease that leads to endothelial dysfunction and disruption of the endothelial barrier, which may underly development of ARDS and proteinuria/AKI.^11,26^ Here, we report increased HPSE activity and HS levels in plasma of COVID-19 patients, which were also associated with severity of the disease.

Several mechanisms are currently proposed to explain pulmonary edema and ARDS in COVID-19. One suggested mechanism focuses on the kallikrein/kinin system, which is involved in the local inflammation and vascular leakage in the lung.^27,28^ Over-activation of the bradykinin pathway may occur due to consumption of angiotensin converting enzyme-2 (ACE2) during viral entry.^28^ Interestingly, endothelial cell surface GAGs, such as HS, regulate activation of bradykinin pathways whereas degradation of HS by bacterial heparinases promotes proteolytic bradykinin generation.^29^ Therefore, increased plasma HPSE activity in COVID-19 patients could contribute to activation of the bradykinin pathway, and subsequently vascular leakage and local inflammation. The renin-angiotensin system also could be involved in endothelial dysfunction in COVID-19 patients.^30^ Increased angiotensin II levels have been reported in COVID-19 patients.^31^ Angiotensin II induces vasoconstriction, inflammation, fibrosis and proliferation, which in turn can cause thrombosis, ARDS and AKI. Importantly, we have previously shown that Angiotensin II is a potent inducer of HPSE expression.^32,33^ Moreover, it is feasible that endothelin-1, one of the downstream mediators activated by angiotensin II^34,35^ is also increased in COVID-19 and it is known that endothelin-1 can induce HPSE expression as well.^36^

Besides the role of HPSE in compromising the endothelial glycocalyx, HPSE and HS fragments play an important role in inflammation.^37^ HPSE can activate macrophages, resulting in secretion of MCP-1, TNF-α and IL-1β, independent of HS-degrading activity.^38^ Released HS fragments also induce a pro-inflammatory response by binding to TLR2 and TLR4.^38,39^ Moreover, cleavage of HS by HPSE releases HS bound molecules, such as chemokines and cytokines, thereby promoting inflammation.^40^ Furthermore, cells exposed to HPSE show an enhanced response to pro-inflammatory cytokines like IFN-γ.^17,41,42^ Interestingly, cytokines such as IL-1β, IL-6, TNF-α and MCP-1 appear to be elevated in COVID-19 patients^43–45^ and also can induce HPSE expression^15^. These data suggest the formation of a HPSE-mediated positive feed forward loop for inflammation in COVID-19. Notably, HPSE appears to have a direct effect in shaping the cytokine milieu, since HPSE deficiency reduces expression of a wide range of cytokines including TNF-α, IL-6, IFN-γ in experimental models.^15^

Potential beneficial effects of prophylactic as well as therapeutic doses of LMWH in COVID-19 patients have been reported.^46–49^ Our data reveal that prophylactic doses of LMWH inhibit HPSE activity in moderately diseased COVID-19 patients, but not in COVID-19 patients in ICU. Therefore, therapeutic LMWH dose instead of prophylactic dose might be required to reduce plasma HPSE activity of COVID-19 patients in ICU. In addition to inhibition of HPSE, LMWH has other non-anticoagulant functions that may be beneficial for patients with COVID-19, such as neutralization of chemokines/cytokines, interference with leukocyte trafficking, neutralization of extracellular cytotoxic histones, neutralization of high molecular weight kinogen, and reduction of viral entry.^29,50-52^

In summary, this cross-sectional study shows that HPSE activity and HS levels are significantly elevated in plasma of COVID-19 patients, which is associated with the severity of COVID-19. Targeting of HPSE activity could be beneficial for the clinical outcome of COVID-19 patients, since it is well established that increased HPSE activity compromises the endothelial glycocalyx and contributes to a pro-inflammatory cytokine milieu. Considering the fact that no specific clinically approved heparanase inhibitors are currently available, prospective studies evaluating the clinical outcome of COVID-19 patients treated with therapeutic doses of LMWH are urgently needed.

## Methods

### Human Samples, demographics and baseline characteristics

This study was performed according to the latest version of the declaration of Helsinki and guidelines for good clinical practice. The local independent ethical committee approved the study protocol (CMO 2020-6344, CMO 2020-6359, CMO 2016-2923). All patients admitted to the Radboud university medical center (Radboudumc) with a PCR-proven SARS-CoV-2 infection was asked for informed consent for participation in this study. After obtaining informed consent, ethylenediaminetetraacetic acid (EDTA) blood was collected and centrifuged for 10 minutes at 2954 *xg* at room temperature (RT), plasma was collected and stored at -80 °C for later analysis. Demographic data, medical history and clinical laboratory measurements were collected from the medical file and processed in encoded form in electronic case report forms using Castor electronic data capture (Castor EDC, Amsterdam, the Netherlands). Plasma was collected from 48 PCR-confirmed COVID-19 patients admitted to the ICU (n= 14) or to designated COVID-19 clinical wards (n= 34). More men than women were included (Table 1). Notably, ICU patients had a significantly higher C-reactive protein concentration than non-ICU patients. The non-ICU patients were further aggregated in those receiving prophylactic LMWH (LMWH+) (in general dalteparin 5000 IU subcutaneously once daily) (n=17) and those receiving either alternative anticoagulation (n=8; vitamin K antagonist n=6, direct oral anticoagulant n=2) or patients for whom the sample collection was performed before initiation of any standard medical intervention (LMWH-) (n=9). Day of sampling was significantly different between ICU and non-ICU groups as well as between LMWH- and LMWH+ groups. The LMWH group had significantly higher median platelet count and D-dimer concentrations compared to LMWH-group, whereas the concentrations of the inflammatory markers CRP and serum ferritin were similar between LMWH+ and LMWH-.

### HPSE activity assay

The activity of HPSE in EDTA plasma was determined by an in-house developed activity assay, which was optimized by the use of recombinant active human HPSE (Bio-techne, Abingdon, United Kingdom, Cat#7570-GH-005). In detail, Nunc maxisorp flat bottom 96 plates (Thermo scientific, Breda, The Netherlands) were coated with 10 ug/ml heparan sulfate from bovine kidney (HSBK) (Sigma-Aldrich, Zwijndrecht, The Netherlands) in optimized HS coating buffer, overnight in a humidified chamber at RT. Subsequently, plates were washed with 0.05% PBS-Tween 20 (Sigma-Aldrich) (PBST) and blocked for minimal 2 hours with 1% bacto-gelatin (Difco laboratories, Detroit, Michigan, USA) in PBS at RT. Plates were washed with PBST, followed by a final washing step with PBS prior to 2 hours incubation at 37°C with 4 times diluted plasma sample in HPSE buffer. Samples were randomly distributed over plates. The HPSE buffer consisted of 50 mM citric acid-sodium citrate (Merck, Zwijndrecht, The Netherlands) buffer supplemented with 50 mM NaCl (Merck), 1 mM CaCl2 (Sigma-Aldrich) and 1 mM DTT (Sigma-Aldrich) at final pH 5.0. Next, plates were washed with PBST and incubated with primary mouse anti-rat IgM HS antibody JM403 (Amsbio, Abingdon, United Kingdom, cat. no. #370730-S, RRID: AB_10890960, 1 μg/ml in PBST) for 1 hour at RT. Subsequently, plates were washed with PBST and incubated with secondary goat anti-mouse IgM HRP antibody (Southern Biotech, Uden, The Netherlands, cat. no. #1020-05, RRID: AB_2794201, dilution 1:10000 in PBST) for 1 hour at RT. Finally, plates were washed with PBST and 3,3′,5,5′-tetramethylbenzidine (TMB) substrate (Invitrogen, Breda, The Netherlands) was added and reaction was stopped by addition of 2 M sulfuric acid, and absorbance was measured at 450 nm. The HPSE activity in plasma was related to a standard curve of recombinant human HPSE in healthy control EDTA plasma.

For the *in vitro* HPSE inhibition experiment with dalteparin (Pfizer, Capelle a/d Ijssel, The Netherlands, Fragmin 12,500 IU/0.5 ml), the HPSE activity was determined using the HPSE activity assay as outlined above. For inhibition studies 0-1 IU/ml dalteparin was used with a constant amount of 150 ng/ml recombinant human HPSE.

### HS competition assay

HS in EDTA plasma samples was quantified by a previously described HS competition assay.^53,54^ Importantly, this assay is specific to HS, therefore the measurement is not affected by the presence of LMWH use. Plates were coated with HSBK and blocked with bacto-gelatin as outlined for the HPSE activity assay. Uncoated plates, blocked with bacto-gelatin, were washed with PBST. The plasma samples were 4 times diluted in PBST containing primary mouse anti-rat IgM HS antibody JM403 (1.3 μg/ml) and incubated for 1 hour at RT. Samples were randomly distributed over plates. Subsequently, the samples were transferred from the uncoated plates to the HSBK-coated plates and incubated for 1 hour at RT. Plates were washed with PBST and incubated with secondary goat anti-mouse IgM HRP antibody for 1 hour at RT. Plates were developed and measured as outlined for the HPSE activity assay. The amount of HS detected in plasma is expressed in arbitrary units since HS from bovine kidney was coated and used to prepare the standard curve.

### Statistical analysis

Values are expressed as mean±SEM. D’Agostino & Pearson normality test was performed to test for normality of data. Significance was determined by Fisher’s exact test to compare categorical variables, by Student’s t-test or Mann Whitney test to compare two groups and by Kruskal-Wallis test followed by Dunn’s test to compare more than two groups using GraphPad Prism V.8.4.2 (La Jolla, USA). P values less than 0.05 were considered as statistically significant.

## Data Availability

Johan van der Vlag has full access to all the data in the study and takes responsibility for the integrity of the data. All data are available in the manuscript.

## Acknowledgements

This study was financially supported by the Radboud university medical center PhD fellow program and consortium grant LSHM16058-SGF (GLYCOTREAT; a collaborative project financed by the PPP allowance made available by Top Sector Life Sciences & Health to the Dutch Kidney Foundation to stimulate public-private partnerships) coordinated by JvdV. MGN was supported by an ERC Advanced grant (#833247) and a Spinoza Grant of the Netherlands Organization for Scientific Research. The graphical abstract was created with BioRender.com.

## Author contributions

BB, CY, QdM and JvdV designed the experiments, analyzed data and wrote the manuscript. BB, CY and MdG performed the experiments. AdN, IG, NAFJ, LABJ, MGN and FLvdV were co-investigators on CMO 2020-6344, which provided COVID-19 patient samples. BB, CY, IJ, NR, RD and JvdV were co-investigators on CMO 2020-6359, which provided COVID-19 patient samples. MK and PP were co-investigators on CMO 2016-2923, which facilitated COVID-19 ICU patient sampling. MLMH created the graphical abstract and wrote the manuscript. TN and LH helped with analysis of the clinical data. JvdV has full access to all the data in the study and takes responsibility for the integrity of the data. All authors critically reviewed and edited the manuscript. BB and CY share first authorship and AdN and IG are co-second authors, listed in alphabetical order.

